# Autism Spectrum Disorder: A Systematic Review on how Diagnosis, Treatment, and Etiology have been considered in Brazil

**DOI:** 10.1101/2020.10.17.20214437

**Authors:** Eduardo Listik, Marcia Listik

## Abstract

Autism spectrum disorder is a group of developmental disorders whose clinical characteristics include socialization impairment, language disability, and unusual behavior. This review aimed to analyze how the Brazilian community researches autism, seeking answers surrounding its treatment, etiology, and diagnosis. The search for publications was based in Elsevier’s Scopus Database in June 2019 and was focused on the study of autism in Brazil or with Brazilian data. We categorized publications on diagnosis, treatment, and etiology. The majority of the publications found after inclusion/exclusion criteria sought to validate and adapt pre-existing scales to the Brazilian Portuguese. Instead, the groups performing these studies have had little background in the biochemical, genetic, or environmental aspects of the disease in the country.

## INTRODUCTION

Autism spectrum disorder (ASD) is a group of developmental disorders that begins, typically, in children below three years. Clinical characteristics of ASD include socialization impairment, language disability, and unusual behavior (e.g., running back and forth, flapping, spinning around) (Park et al. 2016). ASD has become more frequent: in 2012, 1.5% of people in the United States were diagnosed with the disorder (Graf et al. 2017).

There is difficulty in diagnosing ASD as there are no alternative assessments besides clinical evaluation. The health professionals make use of diagnostic tools such as behavior scales in order to facilitate and quantify clinical findings. There are several scales focused on autism, including the ones validated to Brazilian Portuguese such as the Childhood Autism Rating Scale (CARS), the Autism Behavior Checklist (ABC), the Autism Screening Questionnaire (ASQ), the Autism Diagnostic Interview-Revised (ADI-R), the Autistic Traits Assessment Scale (ATA), and the Modified Checklist for Autism in Toddlers (M-CHAT) (Pereira et al. 2008; Marteleto and Pedromonico 2005a; Sato et al. 2009; Becker et al. 2012; Assumpção Jr. et al. 1999; Losapio and Pondé 2008).

This review aimed to analyze how the Brazilian community researches autism, and it seeks answers for its treatment, etiology, and diagnosis.

## METHODS

### Database and Literature Retrieval

The search for publications was conducted in Elsevier’s Scopus Database in June 2019. The precise input was “((TITLE (“autism” AND “Brazil”) OR ABS (“autism” AND “Brazil”)) AND PUBYEAR > 2008”, which translates the terms *autism* and *Brazil* being present in the title or abstract of articles published only from 2009 on.

The search yielded in 67 results, which were screened for eligibility with inclusion and exclusion criteria before further analysis.

### Inclusion Criteria

Articles that focused on the study of autism in Brazil or with Brazilian data were primarily retained in our sample, but we only included researches that had an appeal towards *diagnosis* (either through validated scales, genetic tests, or metabolic biomarkers) and *treatment* (both pharmacological and non-pharmacological approaches) We also considered works that had perspectives on ASD *etiology*.

### Exclusion Criteria

Observational, psychosocial, educational, or behavioral assessment researches were not kept in this study’s sample. Additionally, articles presenting data with autism-like behavior through lesions of the central nervous system or prematurity and articles without methodological controls were also excluded.

## RESULTS

### Study Selection

The search resulted in 67 hits of which only 60 were related to the topic. After a screening of the abstract texts of all these articles, 37 articles were not within our context and were excluded. The remaining 23 were thoroughly read for eligibility assessment, and 10 were excluded: 5 for not presenting methodological controls, 1 for displaying exclusion criteria, 2 for not displaying the inclusion criteria, and 2 for not being fully ASD-related (Figure 1). The remaining 13 articles are described in Table 1.

**Table 1.**
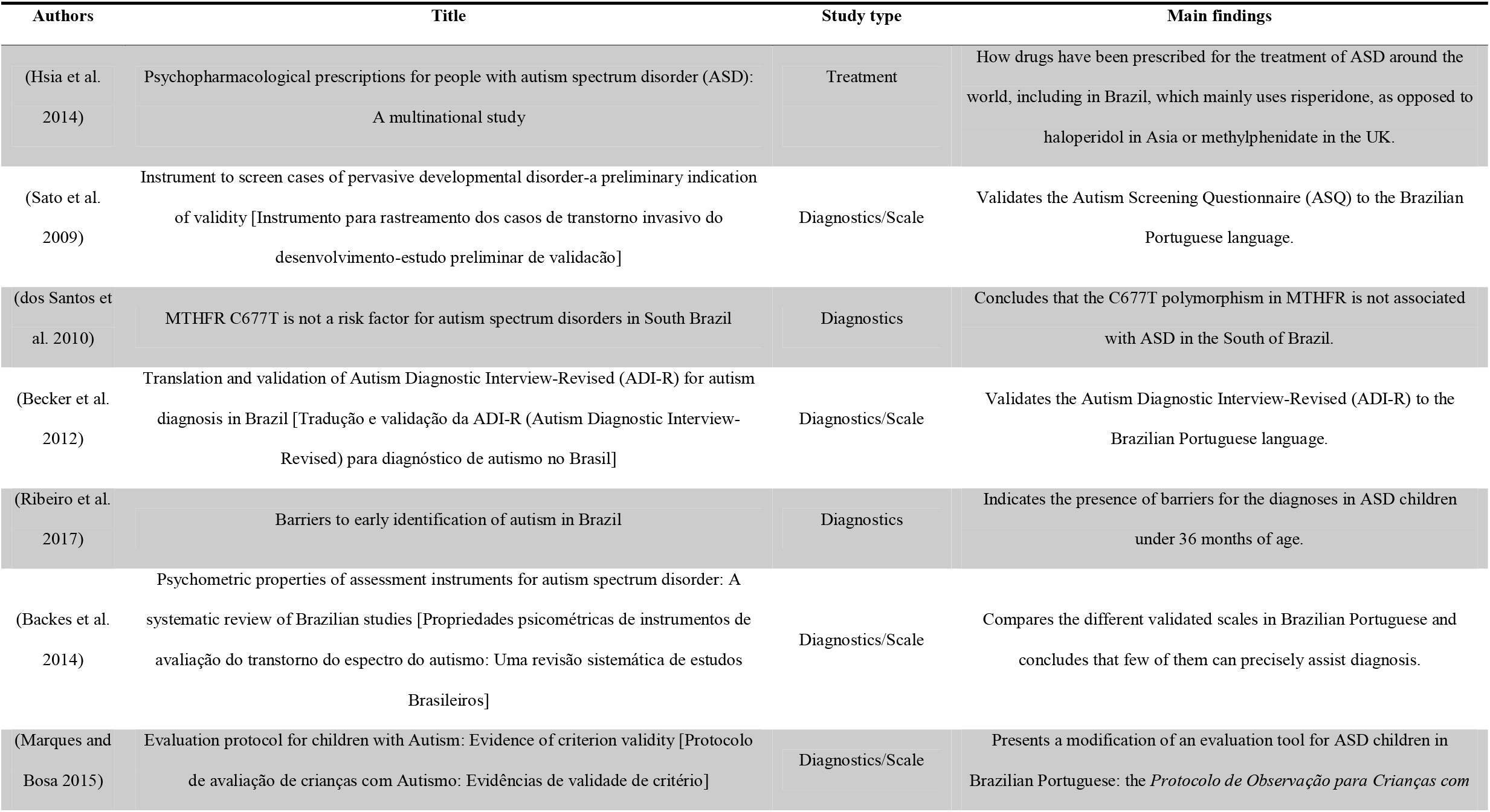
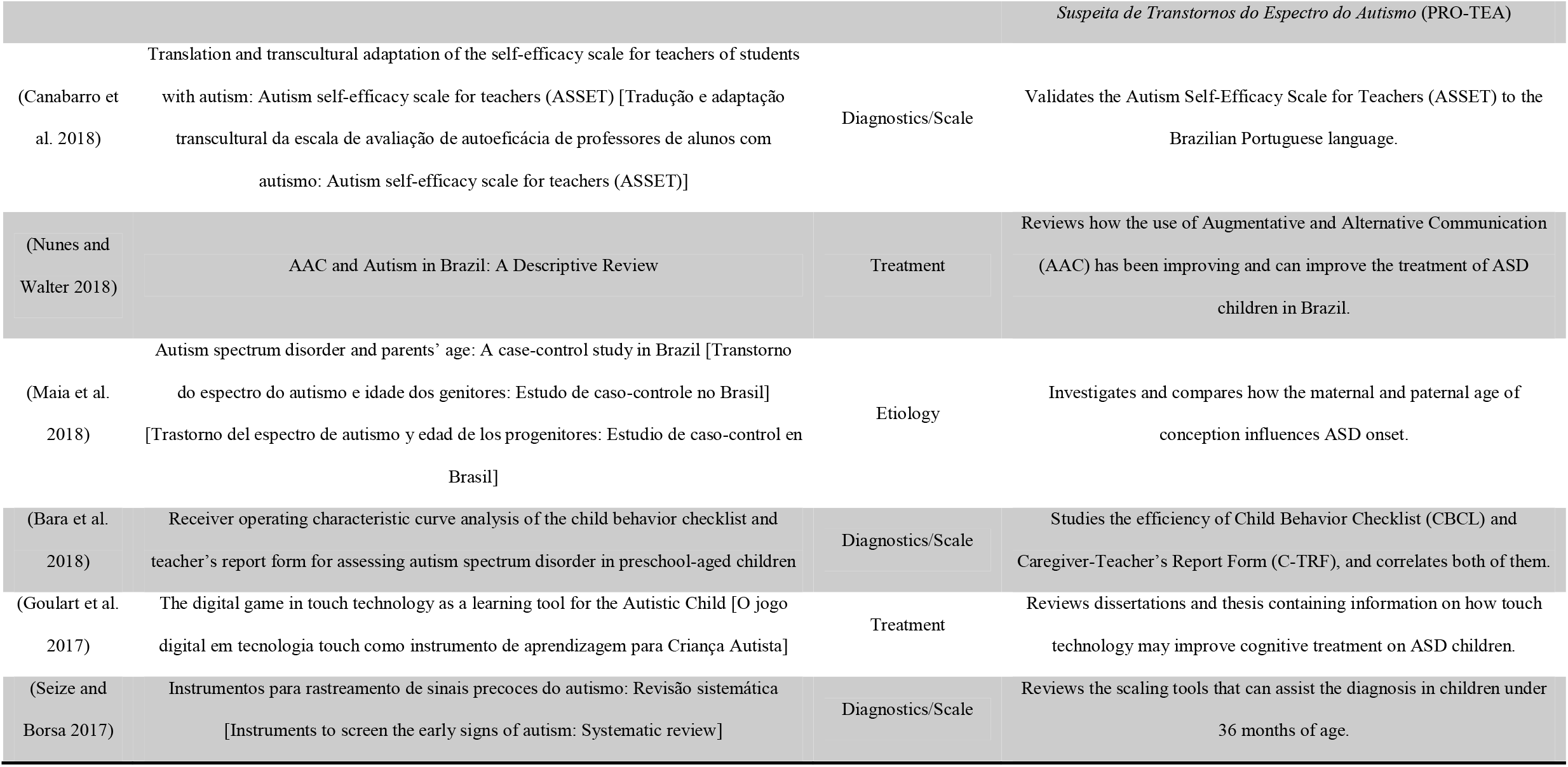
Summary of the findings in this review.

**Figure 1.**
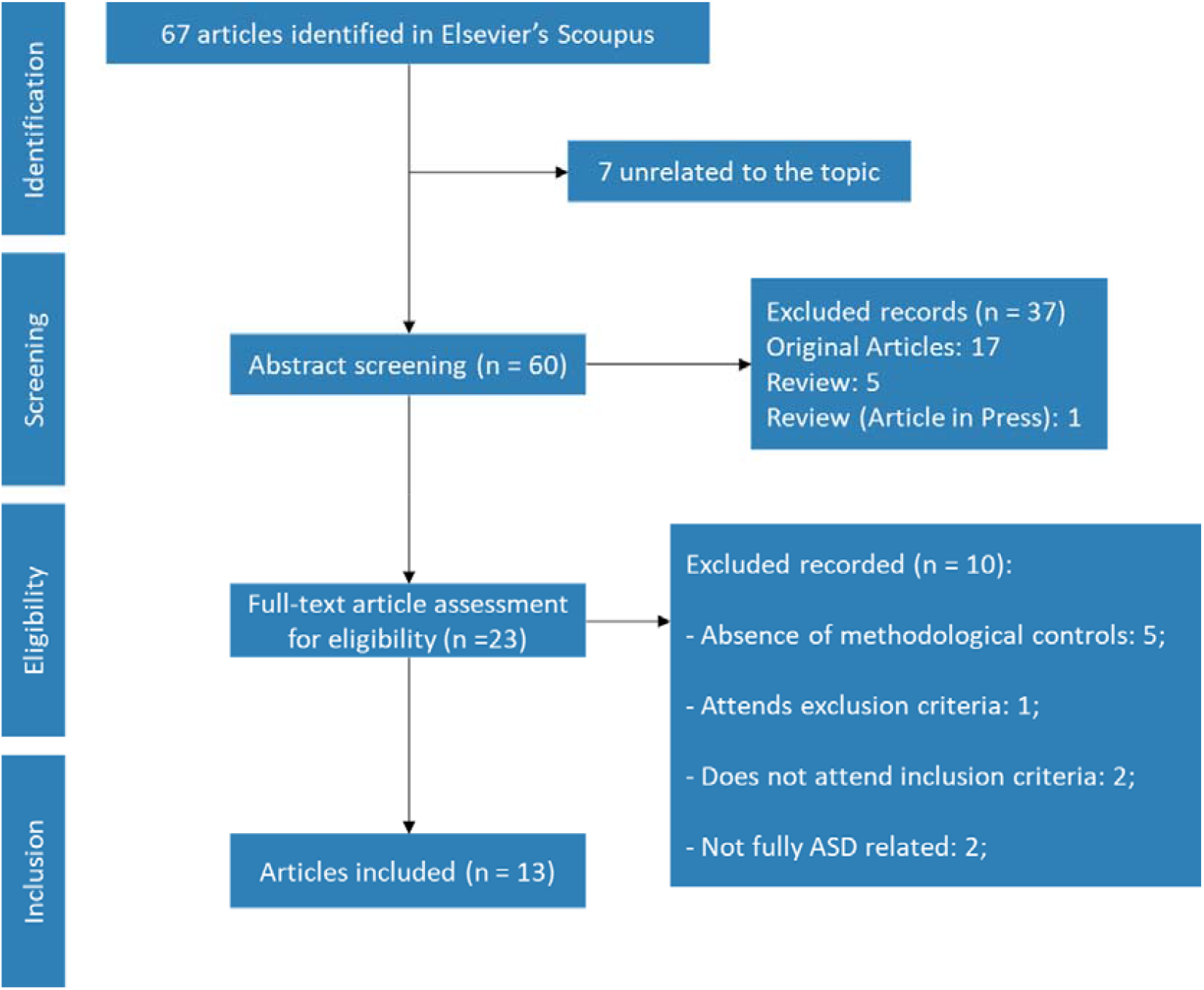
Flow diagram describing the methodology of study selection according to the PRISMA statement.

### Synthesis of Results

#### Diagnosis

Based on our selection, 69% of the articles focused on diagnosis, with 78% of those aimed at validating or adapting a set of scales for autism to Brazilian Portuguese. The other two articles on diagnosis had perspectives on the difficulties of performing a diagnosis of ASD patients of young ages or assessing genetic variation that may (or may not) contribute to autism.

Sato et al. (2009) translated, culturally adapted, and validated the Autism Screening Questionnaire (ASQ) for Brazilian Portuguese (Sato et al. 2009). The validation process included groups of 40 patients, either with a pervasive developmental disorder (PDD), Down Syndrome (DS), or other psychiatric or mental disorders. ASQ has 40 questions, and a score cut-off of 15 indicates a high sensitivity (92.5%) and specificity (95.5%) for PDD alone, distinguishing it from other mental and psychiatric disorders. The study also briefly compares these results from two other previously-validated scales, ATA and ABC (Assumpção Jr. et al. 1999; Marteleto and Pedromonico 2005a), and notes that, unlike ASQ, these scales have been poorly used in the scientific literature; ASQ itself has proven a reliable tool to distinguish PDD from non-PDD cases (DS and other psychiatric or mental disorders).

The Autism Diagnostic Interview-Revised (ADI-R) was first translated and transculturally validated by Becker et al. (2012) (Becker et al. 2012). Their study included 20 patients with autism and 20 patients who did not have the disease but had an intellectual disability. ADI-R contains 93 items, which are presented in an interview to caregivers of possible ASD patients, and it has four scores (A to D), whose cut-offs are 10, 8, 3 and 1, respectively (Lord et al. 1994). The validated scale had high discrimination, revealing a sensitivity and specificity of 100% while *maintaining* the previous cut-off from the English version. The study also mentions that ADI-R is considered a valuable source for autism diagnosis and is frequently used as a clinical instrument (Moss et al. 2008; Gray et al. 2008). It reports that, although 30% of patients with moderate intellectual disability do score in some diagnostic domains of the assay, the study can discretize the groups with confidence (Lord et al. 1994).

The studies found were not only about the validation of scales: Canabarro et al. (2018), for instance, focused solely on translating and adapting the Autism Self-Efficacy Scale for Teachers (ASSET) (Canabarro et al. 2018). This scale possesses 30 items and is intended to inform how teachers may direct themselves towards students with autism (Ruble et al. 2013). The study observes that teachers feel uneasy educating ASD children, primarily due to the variety of symptoms, and it is implied that the use of self-efficacy scales such as ASSET may assist educators in such contexts.

Marques et al. (2015), providing an additional perspective, contributed a study modifying and validating the *Protocolo de Avaliação para Crianças com Suspeita de Transtornos do Espectro do Autismo* (PRO-TEA) (Marques and Bosa 2015). Their subjects were analyzed retrospectively, being divided into three groups of 10 individuals: (i) ASD children, (ii) children with Down syndrome, and (iii) controls. The results demonstrate that the tool may differentiate, among some of its items, ASD children from their controls. Nevertheless, there are still efforts to develop its capacities further, and an investigation is ongoing regarding its sensibility, sensitivity, and other statistics.

Additionally, Bara et al. (2018) promoted more significant insights regarding two different scales for Autism in Brazilian Portuguese that were pending further validation: the Child Behavior Checklist (CBCL) and the Caregiver-Teacher’s Report Form (C-TRF) (Bara et al. 2018). Both scales are questionnaires to be completed by the caregiver/educator of ASD children (from 1.5–18 years) in order to analyze behaviors and indicators of mental disorder (Havdahl et al. 2016; Biederman et al. 2010). Bara et al.’s analysis indicated that CBCL had inferior sensitivity (53.3%) and moderate accuracy, leading to a significant number of false negatives. On the other hand, CBCL demonstrated higher sensitivity (80.6%) and specificity (71.8%). Ultimately, this study acknowledges that the lack of precision in both scales may arise from selection issues with the control group, which contained a variety of behavioral disorders that may have influenced the high false-negative rates.

Backes et al. (2014), on the other hand, performed a very interesting review of the various tools that have been validated for the Brazilian Portuguese and assessed their perspective on ASD (Backes et al. 2014). The review presents six ASD assessment tools that have been validated in Brazilian Portuguese: ABC, ADI-R, ASQ, ATA, CARS, and M-CHAT. It suggests that the M-CHAT was the best-adapted tool and that all six instruments had acceptable internal consistency values. During validation, ASQ, ADI-R, and ABC displayed sensitivity levels above 90%. The authors ultimately name M-CHAT the most reliable tool for ASD assessment and express a desire that more diagnostic instruments be developed to improve clinical practice in Brazil.

Lastly, Seize et al. (2017) is the last study we encountered that focused on how the use of scaling tools may assist diagnostics. This work focuses in particular on instruments that may assist the detection of early signs of autism (prior to 36 months) (Seize and Borsa 2017). The study reports that, although 11 instruments were found for that purpose, only one (M-CHAT) has been validated in Brazilian Portuguese. The authors point out, similarly to Backes & colleagues (2014)(Backes et al. 2014), that this is an alarming landscape: either validation of pre-existing tools or novel tools is needed to assist in the early diagnosis of ASD children, and lack thereof hampers their opportunities to improve the patient clinical perspective among different intervention programs.

Following this context of the scaling tools in ASD, Table 2 summarizes the main statistical characteristics of the scales that were mentioned or present in the studies in this review.

**Table 2.** Overview of statistics regarding validated scales that appeared in our search.

| Study | Scale | Sensitivity<sup>a</sup> | Specificity<sup>b</sup> | Cronbach's α<sup>c</sup> | Median Cohen's $\kappa$ coefficient<sup>d</sup> |
| --- | --- | --- | --- | --- | --- |
| (Sato et al. 2009) | ASQ | 92.5% | 95.5% | 0.895 | Not reported |
| (Becker et al. 2012) | ADI-R | 100% | 100% | 0.967 | 0.824 |
| (Marques and Bosa 2015) | PRO-TEA | Not reported | Not reported | Not reported | Not reported |
| (Bara et al. 2018) | CBCL | 53.3% | Not reported | Not reported | Not reported |
| (Bara et al. 2018) | C-TRF | 80.6% | 71.8% | Not reported | Not reported |
| (Marteleto and Pedromonico 2005b) from (Backes et al. 2014) | ABC | 92.1% | 92.6% | Not reported | Not reported |
| (Assumpção Jr. et al. 1999) from (Backes et al. 2014) | ATA | 96.0% | Not reported | 0.710 | Not reported |
| (Pereira et al. 2008) from (Backes et al. 2014) | CARS | Not reported | Not reported | 0.820 | 0.900 |
| (Castro-Souza 2011) from (Backes et al. 2014) | M-CHAT | 94.0% | 91.0% | 0.950 | Not reported |

<sup>a</sup>Corresponds to how a test correctly identifies the individuals with the disease.
<sup>b</sup>Represents how a test distinguishes the individuals with the disease from those without.
<sup>c</sup>The $\alpha$ represents an internal consistency estimate of the reliability of test scores. Above 0.9 is generally considered acceptable (Streiner et al. 2015).
<sup>d</sup>Verifies how answers agree among different raters. "0" implies no agreement of answers, "1" represents the complete agreement of answers.

Two additional works aimed at the diagnostics of ASD in Brazil were not focused on scaling tools but rather two distinct essential questions. Santos et al. (2010) evaluated a polymorphism (C677T) in the methylenetetrahydrofolate reductase (*MTHFR*) enzyme’s gene to link it potentially with ASD (dos Santos et al. 2010). The study comprised 151 ASD patients and 100 control individuals, who were tested in the South of Brazil, and concluded that this polymorphism alone could not be associated with the disorder. The work also states that several other attempts have been made to draw this connection to *MTHFR*. Nevertheless, the literature presents a handful of mixed conclusions that sometimes indicate that this polymorphism is associated with ASD and other times indicate that it is not (Pasca et al. 2009; Goin-Kochel et al. 2009). As a spectrum, autism is a complex disease, in which biomarkers are complex to establish.

From another perspective, Ribeiro et al. (2017) studied parental dynamics in Brazil, from initial concerns about a possible ASD diagnosis to receiving communication of it from a professional. The group interviewed 19 mothers who had children diagnosed with ASD. The research uncovered that the parents had primary concerns in the first two years of their child’s life, but the concrete diagnosis would only be achieved about three years later. The primary reason cited for the delay was typically the unpleasant or dismissive experiences that the parents would have with health professionals, particularly pediatricians. The conclusion concurs with Seize et

#### Treatment

A small portion of the studies examined aspects of treatment for ASD patients in Brazil.

Hsia et al. (2014) is a study from a Chinese group that investigated the prescription pattern of psychotropic drugs intended for ASD patients’ therapy in an international scope (Hsia et al. 2014). The study analyzed a total of 11 countries, including Brazil, and verified some curious discrepancies. Brazil and Mexico most rarely prescribe psychotropic drugs, as opposed to the highest prescription rates in the USA and Canada. The countries studied mainly utilized risperidone in ASD children, except for the United Kingdom (methylphenidate) and Japan (haloperidol). In Brazil, aside from risperidone (55.7%), other notable drugs in use were quetiapine (7.7%) and carbamazepine (6.7%). It is suggested that some of the diverse prescription profiles may relate to varied aspects of each countries’ health care systems, physician specialties (e. g., pediatricians, psychiatrists, general practitioners), and cultural differences (Murray et al. 2014; Zito et al. 2008).

In another treatment perspective, Nunes and Walter (2018) recently reviewed how Augmentative and Alternative Communication (AAC) may be used as a cognitive development tool for individuals with ASD in Brazil. AAC is, in synthesis, the supplementation of natural speech and writing through aided or unaided strategies in order to promote communication enhancement (Nunes and Walter 2018). The study did conclude that the use of this method is little-explored in Brazil to date: professionals feel ill-prepared to apply it, and the newer models and approaches proposed are not being fully explored as to solidify it as a reproducible method.

Considering these newer strategies, Goulart et al. (2017) proposed to review how the use of touchscreen input devices and their software applications have been employed for Brazilian ASD children (Goulart et al. 2017). It has been demonstrated that these tools have become valuable for teaching academic skills, communication skills, leisure skills, transitioning, and developing employment skills in ASD children around the world (Kagohara et al. 2013). The study revealed that the therapeutic researches of touch technology on ASD children are concentrated in the South of Brazil, which reported evidence that the use of software applications is favorable for ASD children, leading to improvement in behavior, communication, and stereotypical movements.

#### Etiology

Only a single publication within our criteria was found to address the etiology of ASD in Brazil. Maia et al. (2018) investigated, retrospectively, whether the genitor age, either maternal or paternal, could influence the onset of ASD on their children (Maia et al. 2018). They compared 243 ASD patients’ cases with 886 controls and identified that both maternal and paternal ages influence this context, although the former may be more prominent than the latter. The authors argue that both genomic and environmentally-acquired epigenetic alterations become more frequent with aging, and these changes are passed to possible offspring (Puleo et al. 2012; Zhubi et al. 2014).

## DISCUSSION

Our study aimed to review how crucial aspects of ASD have been viewed in Brazil over the last ten years. We searched in the Scopus database and retrieved a total of 67 publications, but after inclusion and exclusion criteria, only 13 were left for analysis, a rather small sample for such a vital theme (Murphy et al. 2016).

As an overall search, for the last ten years worldwide, *autism* yielded 13,501 hits; *autism* and *treatment* 6,748 (Volkmar et al. 2014; Bauman 2010), *autism* and *diagnostic(s)* 3,801 (Constantino and Charman 2016; Fakhoury 2015); and *autism* and *etiology* 1,639 (Risch et al. 1999; Currenti 2010). After our eligibility assessment, there were only 3 publications categorized in *treatment*, 9 in *diagnostics*, and only a single one in *etiology*. This result reveals a very concerning state regarding ASD research in Brazil as very little scientific interest seems to be directed toward this particular field.

What we believe Brazilian ASD research lacks most is an epidemiological assessment of how occurrences may be distributed among region, age, comorbidities, and therapeutics. Such an assessment has yet to be performed for this country, but worldwide, there have been 69 entries for this theme (Wing and Potter 2002; Newschaffer et al. 2007). An epidemiological approach toward ASD patients in Brazil, and a comparison of this profile with other countries, could facilitate possible new insights on causative effects (e. g., environmental, social, and associated disease factors) and an understanding of the disease as a whole (Kim and Leventhal 2015; Duchan and Patel 2012).

The main focus of Brazilian studies for ASD is in diagnostics, mainly in translating, adapting, and validating scaling tools. Although there are additional validated scales in Brazilian Portuguese, they were not included in this review due to their lack of methodological controls. This factor hinders reproducibility of these tools by themselves (Baker and Penny 2016). Additionally, some of the scales discussed in this review lack statistical basis. Many do not report how sensible or specific the validated tool has become after translation, how internally valid it is (Cronbach’s α > 0.9), or even if there is a general agreement by different authorities (i.e., Cohen’s κ) (Streiner et al. 2015). The absence of accurate analysis during validation makes it difficult for professionals to select which tool is fundamentally best for their application (Streiner et al. 2015). Thus, validation strategies in Brazilian studies need improvement. Moreover, many scales do not present cut-off scores to newly-adapted validated versions.

The main article on ASD treatment in Brazil was found to be a multicentric study from a Chinese group; the other two articles reviewed multidisciplinary non-pharmacological treatment (Hsia et al. 2014). There was no discussion of how different physicians (e. g., psychiatrists, neurologists, pediatricians) might proceed in a pharmacological approach in ASD children or adults.(Imran et al. 2011) Moreover, there were no clinical studies to observe a new pharmacological approach or family orientation for this disease (McPheeters et al. 2011; Patterson et al. 2012).

Concerning etiology, only a single work, studying ASD in relation to parental age, was found in a retrospective study. We could not find any publications that traced genetically (Abrahams and Geschwind 2008), environmental (Hallmayer et al. 2011), or metabolic (Ming et al. 2012) causative effects to ASD with Brazilian subjects.

Our study does present some limitations. Firstly, we only used a single database, Scopus, which does not include theses and dissertations. Nevertheless, Scopus features extensive coverage and specificity when compared to other research engines such as Web of Science and Google Scholar (Falagas et al. 2008; Martín-Martín et al. 2018). Additionally, the search terms, although broad, may not have selected all articles that could apply to this systematic review if neither the title nor the abstract contained mention of Brazilian research subjects.

We analyzed studies that focused on autism in Brazil. We have not found an extensive sample and could not verify many groups seeking the etiology of autism in Brazilian subjects. A considerable share of the publications concerned validation of pre-existing scales in Brazilian Portuguese. Instead, the groups lack biochemical, genetics, or environmental perspectives of the disease in the country.

## Data Availability

The datasets generated during the current study are available from the corresponding author on reasonable request

## Notes

### Competing Interest Statement

The authors have declared no competing interest.

### Funding Statement

The authors received no specific funding for this work.

### Author Declarations

This work was exempt from the Universidade Federal de Sao Paulo IRB approval, as it is a review article.

